# “It was like a weird, paralysing thing”: Understanding the development and consequences of emotionally based school non-attendance (EBSNA) through young people’s and parents’ experiences

**DOI:** 10.64898/2026.10.26.26366486

**Authors:** Joanna K. Anderson, Jennifer Saxton, Hayley Gains, Helen Casey, Tamsin J. Ford, Anne-Marie Burn

## Abstract

Emotionally based school non-attendance (EBSNA) is an increasing concern, yet relatively little research has integrated young people’s and parents’ perspectives to understand how attendance difficulties develop and affect young people and their families. This qualitative study explored the development and consequences of EBSNA through the complementary experiences of young people and parents. In-depth semi-structured interviews were conducted with 20 parents and 16 young people aged 11–17 years who had experienced EBSNA. Young people could participate through either verbal or written interviews to support inclusive participation. Data were analysed using the Framework Method, with multiple researchers contributing to coding, charting and interpretation. Three overarching themes were identified: misalignment between young people’s needs and the school environment; becoming unable to attend school; and the wider impact of EBSNA. Across accounts, EBSNA developed gradually through interactions between individual needs and school environments, rather than being attributable to individual characteristics alone. Sensory, social, organisational and academic demands, particularly during the transition to secondary school, combined with unmet needs, bullying, punitive practices and difficulties establishing trusting relationships to progressively undermine psychological safety and belonging. As distress escalated, anxiety increasingly extended beyond specific school situations to school itself, eventually making attendance feel psychologically and physically impossible. Once established, absence created additional barriers to returning through lost confidence, disrupted friendships and concerns about falling behind. Despite substantial educational disruption, young people generally remained motivated to learn. EBSNA also had profound social and family consequences, including isolation, deteriorating parental wellbeing, disrupted family relationships and financial pressures. Findings conceptualise EBSNA as a dynamic socioecological process and highlight the importance of early, individualised responses that address unmet needs and create psychologically safe, inclusive and supportive school environments.

## Introduction

In the UK, persistent school absence (missing 10% or more of school sessions) has increased steadily since 2010 and more than doubled following the COVID-19 pandemic (1, 2). In 2024/25, 18.1% of pupils were persistently absent and 2.4% were severely absent (missing at least 50% of sessions), with substantially higher rates (34.3% and 7.5% respectively) among pupils with an Education, Health and Care Plan (EHCP: a statutory plan for children and young people with special educational needs and disabilities in England) (3).

School attendance problems, including emotionally based school non-attendance (EBSNA), are increasingly recognised as a significant public health concern because of their rising prevalence and association with adverse academic, mental health and socioeconomic outcomes (4). EBSNA refers to a subset of school attendance problems, ranging from intermittent school avoidance to prolonged absence, in which emotional distress is the primary barrier to attendance. Although there is currently no universally agreed definition or operational criteria for identifying EBSNA (5–8), the term is widely used to describe attendance difficulties arising from the dynamic interaction between a young person’s developmental characteristics, emotional wellbeing and school environment. The pathways to EBSNA, its presentation and severity are highly individual and may change over time, highlighting the need for early identification and coordinated, individualised support (4, 9–13).

Estimating the prevalence of EBSNA in the UK is challenging given the lack of an agreed definition, and schools vary in how attendance difficulties driven by emotional distress are recognised and recorded (14, 15). Although UK national policy recommends that absences related to mental health difficulties should generally be recorded as authorised (16), EBSNA-related absence may be classified under a range of authorised and unauthorised absence codes, obscuring its true prevalence. Before the COVID-19 pandemic, approximately 1–2% of school absences were attributed to EBSNA (17). Although concerns about EBSNA have increased since the pandemic, particularly among young people (YP) with mental health difficulties, special educational needs and neurodivergent profiles, evidence of increased prevalence remains limited (18–20). Studies use differing definitions and measures, including broader absence data and qualitative evidence, limiting direct comparisons. A recent UK charity survey found that 28% of secondary school and college students reported avoiding school because of anxiety (21), highlighting the growing scale of anxiety-related attendance difficulties.

Prolonged school absence is linked to a number of adverse outcomes in childhood and adolescence including poor academic attainment (22, 23), school dropout (24), self-harm and suicide (25), increased risk of future mental health problems (26) and impaired social functioning (27). The negative consequences of missing school often extend into adulthood, leading to reduced participation in further and higher education (22, 23), unemployment and economic hardship (28), substance misuse (29), delinquency (30), mental and physical health problems, and early mortality (31). Young people describe deteriorating mental health, disrupted peer relationships, social isolation and uncertainty about their educational futures (32–35). Parents, in turn, report substantial emotional, practical and financial impacts on family life, including chronic stress, reduced employment, financial strain and the demands of caring for and advocating for their child (36–39).

Given the significant adverse consequences of prolonged school absence, understanding the factors that contribute to the development of EBSNA and its impact on YP and their families has become an important research priority. Existing evidence suggests that EBSNA develops through a dynamic interaction between individual, relational and environmental factors rather than a single causal pathway (11, 12). Across studies, anxiety, emotional distress, trauma and neurodevelopmental differences have been identified as individual-level factors increasing susceptibility to EBSNA (5, 12, 40). At the same time, YP consistently describe school environments characterised by overwhelming sensory, social and academic demands, unpredictability, changing routines, bullying, and limited support from school staff as progressively eroding their sense of psychological safety and belonging, eventually making school attendance feel unmanageable (32, 35, 38, 41–43). These experiences seem particularly pronounced among neurodivergent students, who are consistently overrepresented among those experiencing EBSNA (32, 41, 42, 44). In contrast, parents tend to focus on the gradual progression and observable manifestations of distress, describing escalating anxiety, psychosomatic symptoms, emotional exhaustion and withdrawal as reflecting children’s unmet needs within school. Parents describe attendance difficulties developing through the cumulative effects of bullying, academic pressure, sensory overload, feeling misunderstood by school staff and inadequate educational support, with early warning signs often emerging long before attendance deteriorates (34, 45–47).

### The current study

Despite a growing body of qualitative research on EBSNA, qualitative studies integrating the perspectives of both young people and parents remain relatively rare. The present study addresses this gap by exploring the development and consequences of EBSNA through the complementary perspectives of young people and parents.

## Materials and Methods

### Study design

This qualitative study formed part of the Attend Well research programme (attendwell.org.uk), which aims to co-produce a whole-school approach to improving attendance through positive school environments, school connectedness and student mental health. The study involved one-to-one interviews with YP aged 11-18 who experienced EBSNA and their parents/carers. Ethical approval for this work was granted by the University of Cambridge Psychology Ethics Committee (ref: PRE.2025.038).

### Participant recruitment

We used convenience sampling to recruit participants through partner charities, Attendance Support Teams, Local Authority Special Education Needs and Disability (SEND) parent and youth forums, home education networks, SEND Information, Advice and Support Services, and networks of mainstream and alternative provision schools across the UK. Participants were initially invited to complete a quantitative survey. Parents (n = 34) and young people (n = 29) who consented to further contact were subsequently invited to participate in an interview, resulting in interviews with 17 parents and 13 young people. Most young people were recruited through participating parents. An additional three parents and three young people contacted the research team directly and were interviewed before completing the survey. There was no prior relationship between participants and researchers. Participants received information about the study purpose, analysis and dissemination, and had email contact with the research team to discuss and arrange interviews.

### Data collection

Data were collected through in-depth semi-structured online interviews, conducted by JKA with support from HG, between October 2025 and June 2026. Separate but parallel topic guides for parents and YP were developed and refined following review by the Attend Well Parent Advisory Group and three YP (Appendix 1). The guides explored the development of EBSNA, the role of mental health and neurodivergence, school experiences, relationships with peers and staff, factors contributing to attendance difficulties, experiences of support, and views on what facilitates successful re-engagement with education. Parents reflected on their child’s experiences, whereas YP discussed their own.

We offered YP a choice to participate in either an online semi-structured interview or a written interview. Nine completed verbal interviews and seven chose the written format because they did not feel comfortable speaking directly with a researcher. The written interview was a shortened version of the topic guide comprising open-ended questions covering the same key areas, enabling inclusive participation while maintaining consistency across interview formats (Appendix 2). All verbal interviews were audio-recorded and transcribed using Microsoft Teams. Following each interview, the transcript was downloaded, de-identified, and checked against the audio recording for accuracy by a member of the research team. Interviews lasted between 27 and 63 minutes, with most lasting approximately one hour. No notes were taken during interviews.

### Data analysis

All members of the research team had experience conducting qualitative research and had completed relevant methodological training. AMB has extensive expertise in qualitative research, including teaching qualitative methods and contributing to multiple qualitative studies, and provided methodological oversight throughout the analysis.

We analysed the data using the Framework Method of thematic analysis (48, 49). Following familiarisation with the transcripts, the senior authors developed an initial analytic framework informed by the study research questions and interview topic guides. To enhance rigour, members of the research team independently coded two overlapping transcripts using the preliminary framework. They applied both deductive and inductive coding and proposed additional codes to refine and expand the framework, ensuring that it adequately captured the breadth of participants’ experiences.

We entered the revised analytic framework into NVivo 15 (50) and applied it across all transcripts. Five researchers independently coded the data, applying the framework deductively while continuing to code inductively to identify concepts not represented in the initial framework. We met regularly throughout the analysis to discuss coding, resolve discrepancies and iteratively refine the analytic framework. An experienced qualitative researcher (AMB) oversaw the analytic process.

We exported a framework matrix from NVivo into Microsoft Excel, organising the data by cases (rows) and codes/themes (columns). The matrix contained the verbatim extracts assigned to each code. Members of the research team then charted the data by producing summaries of the extracts and recording analytic notes to facilitate systematic comparison across participants. To promote consistency, all researchers independently charted the same two transcripts and compared their interpretations before completing the remaining dataset. We then synthesised the charted data to identify patterns, relationships and overarching interpretations across cases and themes, which we organised into the final themes and subthemes. Finally, we presented the final thematic framework to Attend Well’s two advisory groups: the Young People’s Advisory Group and the combined Parent and Professional Advisory Group. Their feedback informed the interpretation and refinement of the findings. The final thematic framework, including detailed descriptions of each code and subtheme, is presented in Appendix 3, with a simplified overview provided in Figure 1.

**Figure 1:**
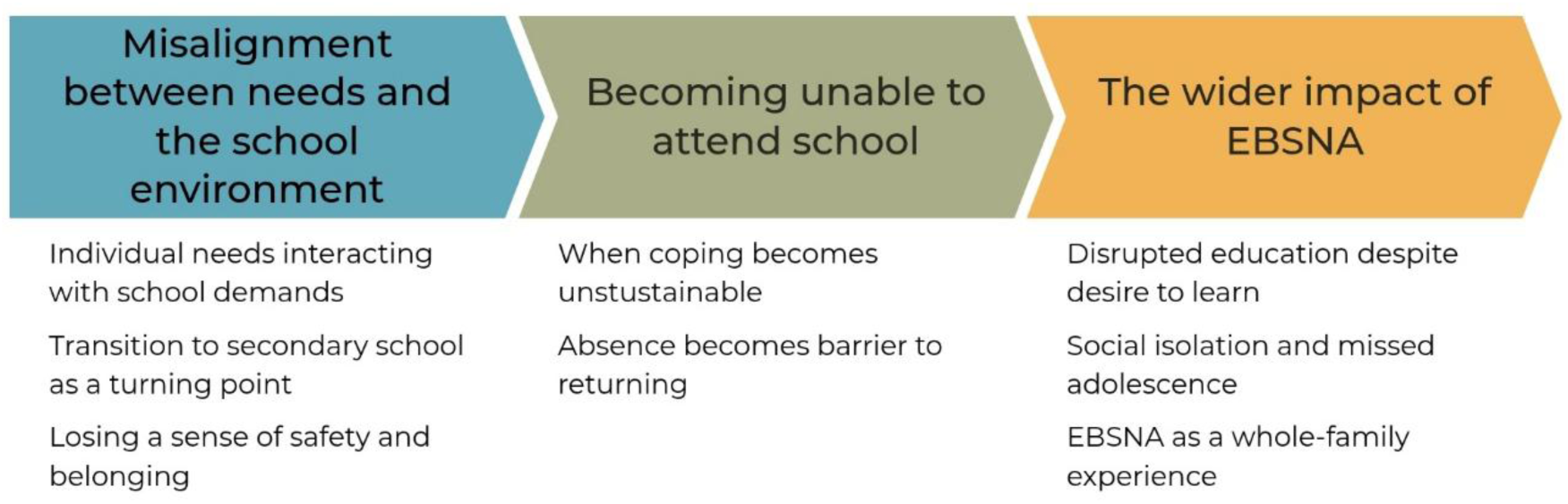
Final thematic framework.

### Research team

The research team comprised five female researchers from one English university, primarily from White, middle socio-economic backgrounds. We all had experience in qualitative and applied mental health research.

We recognised that our social, educational and professional backgrounds may have shaped both data collection and interpretation. The study lead was close in age to many participants and brought relevant lived experience to the study, which may have facilitated rapport, empathy and openness during interviews. At the same time, we remained mindful of how our shared experiences and assumptions could influence interpretation of the data. Throughout the study, we engaged in regular reflexive discussions to critically examine our assumptions, consider alternative interpretations and enhance the credibility of the findings.

## Results

Reporting of the study followed the Consolidated Criteria for Reporting Qualitative Research (COREQ) guidelines (51) (Appendix 4).

### Characterises of participants

Twenty parents and 16 YP participated in the interviews. Parent participants had a mean age of 49.6 years (range 32–57), were all female and predominantly White British (90%). The YP discussed in parent interviews had a mean age of 13.7 years (range 11–18); half had an EHCP, 90% were described as neurodivergent, and 75% were reported to have a current mental health difficulty. The YP interview sample had a mean age of 13.7 years (range 11–17) and comprised predominantly female (69%) and White British (88%) participants. Half attended mainstream secondary schools or colleges, while almost one-third attended specialist provision. Most were in full-time education (94%), 75% had an EHCP, 63% identified as neurodivergent, and 56% reported a diagnosed mental health difficulty. For detailed characteristics of participants see Table 1 below.

**Table 1:** Participant characteristics.

| Characteristic | Parents interviewed (N = 20) | YP discussed by parents (N = 20) | YP interviewed (N = 16) |
| --- | --- | --- | --- |
| **Age, years, mean (range)** | 49.6 (32–57) | 13.7 (11–18) | 13.7 (11–17) |
| **Sex registered at birth** |  |  |  |
| *Female* | 20 (100) | 9 (45) | 11 (69) |
| *Male* | 0 (0) | 11 (55) | 5 (31) |
| **Gender identity matches sex registered at birth** | NC |  |  |
| *Yes* | — | 18 (90) | 14 (88) |
| No | — | 2 (10) | 2 (13) |
| **Sexual orientation** | NC | NC |  |
| *Heterosexual (straight)* | — | — | 8 (50) |
| *Gay* | — | — | 4 (25) |
| *Prefer not to say* | — | — | 4 (25) |
| **Ethnicity** |  |  |  |
| *White British* | 18 (90) | 16 (80) | 14 (88) |
| *Other ethnic background* | 2 (10) | 4 (20) | 2 (13) |
| **Geographical location** |  |  |  |
| *London* | 4 (20) | 4 (20) | 2 (13) |
| *Midlands/East Midlands* | 3 (15) | 3 (15) | 3 (19) |
| *East of England* | 8 (40) | 8 (40) | 6 (38) |
| *South West* | 5 (25) | 5 (25) | 3 (19) |
| *South East* | 0 (0) | 0 (0) | 2 (13) |
| **Current educational placement** | NC |  |  |
| *Mainstream secondary school/college* | — | 14 (70) | 8 (50) |
| *Specialist secondary school* | — | 3 (15) | 5 (31) |
| *Virtual school, home education or other* | — | 3 (15) | 3 (19) |
| **Currently in full-time education** | NC | NC |  |
| *Yes* | — | — | 15 (94) |
| *No* | — | — | 1 (6) |
| **Eligible for free school meals** | NC |  |  |
| *Yes* | — | 3 (15) | 2 (13) |
| *No* | — | 17 (85) | 14 (88) |
| **Education, Health and Care Plan (EHCP)** | NC |  |  |
| *Yes* | — | 10 (50) | 12 (75) |
| *No* | — | 10 (50) | 3 (19) |
| *Don't know* | — | 0 (0) | 1 (6) |
| **Neurodivergent** | NC |  |  |
| *Yes* | — | 18 (90) | 10 (63) |
| *No* | — | 2 (10) | 4 (25) |
| *Don't know* | — | 0 (0) | 2 (13) |
| **Current/diagnosed mental health difficulty** |  |  |  |
| *Yes* | 13 (65) | 15 (75) | 9 (56) |
| *No* | 7 (35) | — | 4 (25) |
| *Don't know* | — | 5 (25) | 3 (19) |
| **Long-term physical health condition** | NC |  |  |
| *Yes* | — | 7 (35) | 7 (44) |
| *No* | — | 13 (65) | 7 (44) |
| *Don't know* | — | 0 (0) | 2 (13) |
| **Highest educational qualification** |  | NC | NC |
| *A-levels* | 1 (5) | — | — |
| *College or university degree* | 13 (65) | — | — |
| *Postgraduate degree* | 6 (30) | — | — |
| **Employment status** |  | NC | NC |
| *Full-time employment* | 8 (40) | — | — |
| *Part-time or not currently working* | 12 (60) | — | — |
| **Living with** | NC | NC |  |
| *Both parents* | — | — | 11 (69) |
| *One parent or other caregiver* | — | — | 5 (31) |
| **Parent has a mental health condition** | NC | NC |  |
| *Yes* | — | — | 8 (50) |
| No | — | — | 4 (25) |
| Don't know | — | — | 4 (25) |
| **Financial worries** |  | NC |  |
| Often | 6 (30) | — | 3 (19) |
| Sometimes | 6 (30) | — | 3 (19) |
| Never or hardly ever | 8 (40) | — | 9 (56) |
| Prefer not to say | 0 (0) | — | 1 (6) |

Values are n (%) unless otherwise indicated. Percentages may not sum to 100 due to rounding. YP = young people; EHCP = Education, Health and Care Plan; NC = not collected.

Three overarching themes were identified across the young person and parent accounts, reflecting both shared experiences and areas of differing emphasis between the two groups. All three themes were informed by data from both young people and parents, although the relative contribution of each group varied across themes and subthemes. The themes are presented below with illustrative quotations from both groups. Quotations are identified using Y for young person and P for parent.

### Theme 1: Misalignment between needs and the school environment

### Individual needs interacting with school demands

Across YP’s and parents’ accounts, EBSNA developed through an increasing mismatch between YP’s needs and the school environment. Participants emphasised that emotional, physical and neurodevelopmental conditions increased vulnerability but did not inevitably lead to EBSNA. Instead, attendance difficulties arose when school environments failed to accommodate individual needs.

Neurodevelopmental differences were commonly discussed by both YP and parents. YP with autism, ADHD and sensory differences did not view their diagnoses as sufficient explanations for their attendance difficulties. Instead, they described an interaction between their needs and environments characterised by sensory stimulation, changing routines, and complex organisational and social expectations. One participant explained:

> *“… if it’s assessment week… I’ll have to move sets and then I’ll have to meet new people again.”* (Y003)

Parents strongly supported these accounts as they described everyday aspects of school life becoming overwhelming for children who depended on predictability and consistency. One parent explained:

> *“Certain things that she just cannot tolerate, which makes attending school really, really difficult… she can’t tolerate any kind of change of classroom… she needs everything to be the same, predictable and that’s kind of in the context of her autism diagnosis.”* (P023)

Both YP and parents frequently described the sensory overwhelming nature of mainstream school environments. Crowded corridors, noisy classrooms, busy lunch halls, bright lighting, strong smells and close physical proximity to others were frequently described as persistent sources of distress. One young person summarised the difficulty as *“the whole environment… the noise, the people”* (Y001), while another avoided travelling on the school bus because there were “*loads of crowds”* and *“all these people touching you”* (Y003). One young person reflected:

> *“I’m one of those people who finds the environment really tricky… the canteen is really small and noisy, there’s nowhere else to go… the lighting is really bright… it’s spray, perfume, food…”* (Y012)

Parents described the same environmental demands but provided further insight into the impact they had on some YP. One parent explained:

> *“The bright lights, the noise, the whole business of just school life… completely threw him, he went into almost like shut down. He would stand in a corridor and cry or shake and his brain will go completely into freeze and wouldn’t know where he was going, what he’s doing.”* (P030)

YP and parents agreed that strict uniform policies often resulted in unnecessary discomfort, particularly for YP with sensory sensitivities. As one parent explained, *“you’ve got your tie, your blazer, your jumper… that’s quite a lot for her”* (P020). YP similarly described uncomfortable uniforms as *“another pointless barrier”* (Y007), with one recalling being required to wear a blazer in 31°C heat (Y012) and others described wearing a unform “*24/7*“ as *“a massive struggle”* that created *“unnecessary discomfort”* (Y020).

A number of parents described children developing physical symptoms as the demands of the school environment increasingly exceeded their capacity to cope. Many reflected that it was only in retrospect that they recognised these as early signs of mounting distress rather than isolated difficulties. One parent reflected:

> *“There was a lot of sickness as well. A lot of ‘my throat hurts, my stomach hurts’… looking back, there were so many clear signs here.”* (P034)

Several families initially sought medical explanations for these symptoms. One parent described repeated hospital appointments before recognising the connection between their child’s physical distress and school:

> *“He was having physical symptoms. Abdominal cramps. He’s been backwards and forwards to hospital under paediatricians…”* (P030)

In contrast to parents’ emphasis on physical symptoms, YP more often described the emotional experience of becoming unable to cope with school. Many talked about anxiety, low mood and emotional exhaustion that gradually intensified over time, making attendance feel psychologically and physically impossible rather than a matter of choice. One young person explained:

> *“I do remember like this kind of blocker in my head just appearing… I finally woke up and was like, ‘You know what, I’m just not going to go… I’m just going to stop fighting.’ It was waiting to happen.” (Y010)*

Parents also highlighted YP’s sustained emotional effort they made to remain in school. Many described children masking their distress, monitoring their behaviour and attempting to meet expectations throughout the day while appearing able to cope. One parent characterised this as a state of chronic stress:

> *“You’re going in holding your breath and then you’re holding your breath the whole day and you don’t get to breathe till you get home. It’s like just chronic stress… for a neurodivergent person.”* (P019)

Together, these accounts suggest that EBSNA emerged through a growing mismatch between YP’s needs and school environments, with parents extending YP’s accounts by highlighting physical symptoms of anxiety and cumulation of distress as well as YP’s attempts at masking them.

### Transition to secondary school as a turning point

Across YP’s and parents’ accounts, the transition from primary to secondary school often represented a critical point in the development of EBSNA. Although some YP had already experienced emotional or attendance difficulties in primary school, previously manageable challenges became overwhelming at this transition. Rather than a single precipitating event, it involved a simultaneous increase in academic, organisational and social demands alongside the loss of familiar routines, friendships and environments. For many YP, these changes marked the point at which attendance rapidly deteriorated. One participant reflected:

> *“I found it very hard and confusing… I do think Year 7 was much worse… I hated school so much that I started staying up until about 5 a.m. every single day so that I could sleep through the day.”* (Y015)

YP consistently described secondary school as fundamentally different from primary school. Larger school sites, unfamiliar teachers and peers, constantly changing classrooms and routines, and greater expectations for navigating it all independently created a sense of uncertainty and loss of control. One participant described moving *“from the baby school to the school where the Year 11s are adults”* (Y002), reflecting not only the physical size of secondary school but also the expectation to function independently within a far more complex environment. Others recalled struggling to manage multiple online systems, timetables and school procedures, feeling they were expected to understand how secondary school operated without sufficient guidance or support.

Parents’ accounts corroborated these experiences, describing how children who relied on routine, predictability or adult support found the transition particularly challenging. They reflected that the increased complexity and expectations for independence often exceeded their child’s capacity to cope from the outset. As one parent explained:

> *“They’re expected to know where to be, when to get there, how to get there, find their own way around school. He had these massive classes, these busy, busy, busy corridors.”* (P012)

Another parent reflected that, after moving from a much smaller primary school, her son became *“one tiny speck in a giant school”* (P034), while another described *“the enormity of accessing secondary school and absolutely everything that’s expected and involved… he couldn’t do.”* (P012).

Beyond navigating the physical environment, participants described the transition as disrupting the familiarity and predictability that had previously enabled many YP to cope. YP recalled leaving small primary schools where they knew the environment, routines, staff and peers—and were themselves well known—to much larger secondary schools where they often felt anonymous, unsupported and uncertain about how to navigate everyday school life. One young person explained:

> *“I get anxious because I have to meet new people and then the classroom will change. And then I have to go and find the different classrooms.” (Y005)*

Both YP and parents described how social demands became a challenge in secondary school. YP reflected on leaving established friendship groups and struggling to build new relationships, while parents explained how unstructured social situations and increasingly complex interactions were challenging, particularly for neurodivergent children. As one parent reflected:

> *“…moving up to secondary school, … all of a sudden, there’s all that free time that can only be filled by social communication. And he only wants to talk about his special interests… he’s not interested in chit chat.”* (P027)

Similarly, another parent described her child as living *“in absolute fear”* that peers would ask them to contribute to conversations because they struggled to follow what others were discussing (P019).

Although both groups identified secondary school as a critical turning point, parents introduced an additional factor that was largely absent from YP’s accounts - inadequate transition planning. Several participants described poor communication between primary and secondary schools, meaning that YP’s identified needs and established adjustments were not communicated to their new school. One parent explained that despite them sharing information about her daughter’s attendance difficulties, previous support and CAMHS involvement before the move to secondary school, *“no work was done to transition her”* (P029). As they reflected:

> *“I don’t think anyone ever read that. And the SENCO that was there in her Year 7, I never got to meet her. I emailed. I never heard back.”* (P029)

Overall, both YP and parents described the transition to secondary school as a tipping point at which existing vulnerabilities became increasingly difficult to manage. YP described navigating unfamiliar environments while dealing with increasing expectations, whereas parents highlighted how insufficient preparation and inadequate transition planning further compounded these challenges.

### Losing a sense of safety and belonging

Both YP and parents described EBSNA developing through a gradual erosion of psychological safety and belonging at school. Repeated experiences of peer difficulties, feeling misunderstood by adults, harsh disciplinary practices and unmet needs left YP feeling increasingly unsafe and disconnected from school.

YP frequently described bullying, social exclusion and difficulties developing or maintaining friendships, particularly following the transition to secondary school. One participant described how changing tutor groups to escape bullying inadvertently increased their isolation, as everyone else already knew one another, leaving them feeling *“a bit like an outsider.”* (Y010). Another described being *“scared of walking around”* school because they expected other pupils to *“shout and laugh”* at them (Y005).

Parents described similar difficulties forming friendships, feeling different from peers and repeated experiences of rejection. As one parent explained:

> *“Everything started when he felt like he wasn’t being well liked in the school. (…) he would tell me things like ‘I don’t feel like I belong there. I don’t feel comfortable in the school. I don’t have friends.’”* (P002)

A few YP described experiences of persistent and severe bullying that was inadequately addressed by schools. One participant recalled:

> *“Only one kid ever got punished… I had kids throwing stuff at me… paper clips, pencils, pens, nails at some point… They did nothing… and I think that just really allowed it to start to happen more.”* (Y019)

Parents echoed these accounts but also highlighted experiences of discrimination. Some described children being labelled as *“weird”* because they *“found communication very difficult”* (P030), while others reported prejudice linked to the intersection of neurodivergence and race. As one parent reflected, their daughter’s experiences could not be understood through autism alone:

> *“…when she’s quiet people think she’s being rude, when she speaks up, they think she’s being aggressive. So it feels like she can’t win. So as a Black autistic girl, I feel she’s caught in-between and her struggles get overlooked.”* (P001)

Alongside peer relationships, both YP and parents consistently described feeling misunderstood by school staff. YP frequently reported that teachers interpreted distress, sensory overload or attempts to self-regulate as deliberate misbehaviour rather than recognising them as expressions of unmet need. One participant with ADHD described repeatedly being told to *“sit still, sit quiet, stop moving”* because teachers failed to recognise these behaviours as attempts to self-regulate (Y012). Parents’ accounts corroborated these experiences, explaining that many children masked their distress or appeared to cope well in school, leading staff to underestimate the extent of their difficulties. As one parent explained:

> *“The number of times I’ve heard that he’s fine, but actually he was dissociating… counting lights on the ceiling as a coping strategy.”* (P021)

Although not reflected in parents’ accounts, several YP described feeling judged, dismissed or bullied by school staff. Rather than recognising distress or unmet needs, some teachers responded with scepticism, criticism or confrontation, undermining YP’s trust in school. One participant recalled repeated negative interactions with a member of the SEND department, explaining:

> *“This teacher was bullying autistic children… she started implying that I was lying about being able to walk. I talked to my mother and… that’s what eventually pulled me away from school.”* (Y019).

Parents additionally described how disrupted or inconsistent relationships with trusted adults contributed to attendance difficulties emerging or escalating. One parent recalled that her son had begun to form an attachment to a new teacher, but *“that very night”* learned she was going on maternity leave, which she described as *“the beginning of his school avoidance”* (P021). Another parent reflected that although.

> *“…he was allocated a TA to try and build a relationship with him… she couldn’t do it consistently and he just wasn’t able to build a relationship with them”* (P027).

Most YP also described wider school cultures that prioritised compliance over wellbeing. Rigid behaviour policies, public reprimands, isolation rooms and sanctions for minor infractions commonly increased anxiety. One participant questioned the value of punitive disciplinary approaches, explaining:

> *“If someone has a behavioural issue, shaming them… isn’t actually going to help… they were put in isolation rooms all day… they didn’t learn anything.” (Y007)*

Parents highlighted how behavioural policies and inflexible school routines undermined children’s sense of safety, particularly for neurodivergent YP. They described restricted access to toilets, rigid expectations around organisation and classroom behaviour, and limited flexibility to accommodate children’s specific needs. As one parent explained:

> *“It’s the behaviour policies of the school … be prepared, eye contact, put your hand up, ask questions, be organised, always have all your equipment with you. Every single one of them behaviour policies directly discriminate anybody with any disabilities”* (P020).

Across both groups, EBSNA developed as school experiences gradually undermined YP’s sense of psychological safety. While YP described schools as socially threatening and emotionally unsafe, parents highlighted the importance of trusting adult relationships and the cumulative impact of unmet needs.

### Theme 2: Becoming unable to attend school

#### When coping became unsustainable

Across both datasets, participants described how escalating anxiety gradually exceeded YP’s capacity to cope, culminating in deterioration in mental health and functioning. What began as anxiety about specific situations progressively extended to anticipating school itself, making everything associated with being in school feel psychologically overwhelming.

YP frequently described severe anxiety that extended well beyond the school day, with constant worries about school-related demands, forgetting equipment, receiving detentions or coping with assessments. One participant described becoming *“very annoyed and scared”* every Sunday because of the thought of returning to school, while assessment weeks routinely left them unable to sleep until *“like 2.00 AM”* (Y005). Others described anxiety becoming so overwhelming that they were physically unable to prepare for school.

> *“The anxiety would not let me… it was like a weird, paralysing thing. I physically could not go there. I couldn’t put on my uniform or anything. I couldn’t do anything that would lead to me going to school.”* (Y007)

Parents’ accounts closely mirrored these experiences, describing anxiety extending progressively further back from the school day. What initially appeared as reluctance to attend in the morning gradually became anxiety on waking, the evening before and, eventually, several days in advance, illustrating how anticipation of school itself had become overwhelming.

> *“It started off with not wanting to go in the mornings, but then it kind of moves to, as soon as they wake up, don’t want to go to school and then it moved to the night before and then it was kind of even teatime the night before they started to worry about.”* (P020)

As anxiety intensified, both groups described school becoming predominantly associated with fear. YP explained that fear no longer centred on individual lessons or events but on entering the school environment itself, while parents described children increasingly viewing school as an unsafe place despite continued efforts to encourage attendance. One parent reflected that her son experienced school as *“a hellhole… torture… and a prison”* (P024).

Participants described this escalating distress as accompanied by a progressive deterioration in mental health and functioning. Their children became increasingly isolated, no longer leaving the house, refusing to eat or communicate, while YP spoke of becoming emotionally numb, withdrawing from others and struggling to express how they were feeling. One parent explained:

> *“…There would be a knock at the door, and he would go into a panic. He would sit all day with a blanket over his head and not talk to anyone and not eat.”* (P005)

Both groups also described increasingly severe mental health difficulties. YP recounted periods of depression, intrusive thoughts, self-harm and suicidal ideation:

> *“I was really depressed at the time… I was getting loads of suicidal thoughts… then I started to get loads of intrusive thoughts as well… I was then non-verbal, I was self-harming…“*(Y012)

Parents corroborated these accounts, recalling children expressing that they no longer wanted to live, engaging in self-harming behaviours or requiring constant supervision to keep them safe. One parent explained that they had to *“hide all the knives in the house”* because their son *“would rather be dead and not be here”* (P005).

For some families, this deterioration culminated in an acute mental health crisis that fundamentally changed how school attendance was viewed. Parents described reaching a point when insisting on attendance no longer felt safe or appropriate. One parent recalled that their son *“basically had a mental health breakdown”*, attempted to end his life, and was no longer able to attend school (P018), while another described how their priorities shifted after watching her son *“put a blanket over his head and… sob”,* explaining, *“I looked at him and I thought, I’m not going to put this boy through this again”* (P024).

Together, these accounts describe a progression from escalating anxiety to profound psychological withdrawal and, for some, mental health crisis. YP conveyed the internal experience of emotional exhaustion, depression and trauma, whereas parents illuminated the gradual deterioration they witnessed, including autistic burnout, withdrawal from everyday life and suicide attempts. Across both perspectives, EBSNA became entrenched when the sustained effort of coping with school demands exceeded YP’s psychological resources.

### Absence becomes barrier to returning

Across both YP’s and parents’ accounts, EBSNA became increasingly self-perpetuating over time. Although attendance difficulties often began in response to overwhelming school experiences, participants consistently described absence itself becoming an additional barrier to returning. As time away from school increased, YP lost confidence in their ability to reengage, while anxiety about returning continued to escalate.

For some YP, attendance was initially disrupted by a physical illness, after which returning became increasingly difficult. As one participant explained, *“I had a big surgery around Christmas and that just set me back completely and then just basically didn’t go in the rest of the year”* (Y002). They later reflected that prolonged absence had become the main obstacle because *“it’s now more just difficult going back because… it’s been such a long time”* (Y002).

Others similarly described the original reasons for struggling to attend gradually becoming overshadowed by fears about reintegration. As school routines continued without them and classmates progressed, returning came to feel increasingly daunting. One young person explained:

> *“It’s just been difficult to get that going… because I haven’t done it for, what, like four years, five years… It just seems like a bigger thing than it probably is. I probably get in and I’d be fine.”* (Y015)

Many YP described the loss of friendships during prolonged absence as a major barrier to returning to school. Many worried that friendship groups had changed while they were away and felt increasingly anxious about returning to a school where they no longer knew anyone. One participant explained that *“I don’t have any friends at the moment really”* making returning *“more difficult because I don’t know anybody”* (Y002). Others described feeling isolated during the school day, with one participant explaining, *“I sit by myself in a room at break and lunch”* (Y012) because they had no one to sit with.

Parents described the same process, observing children becoming increasingly anxious about returning after prolonged absence. Many worried about falling behind academically, being judged by teachers or peers, or simply having to explain their absence. One parent explained that her son constantly worried about *“falling behind or people judging him for not being there”* (P002), while another described their child as *“desperately afraid of getting left behind”* (P018).

Together, these accounts suggest that prolonged absence became a barrier to reengagement. As confidence declined and school became increasingly unfamiliar, returning felt progressively more overwhelming, requiring support that addressed not only the original sources of distress but also the new barriers created through absence.

### Theme 3: The wider impact of EBSNA

#### Disrupted education despite desire to learn

Across both YP’s and parents’ accounts, EBSNA had substantial educational consequences, disrupting learning, limiting access to education and reducing opportunities to achieve academic potential. However, participants in both groups consistently emphasised that this reflected an inability to attend school rather than a lack of motivation to learn.

Many YP described receiving little or no structured educational support during periods of absence. Several explained that schools provided minimal work or expected them to learn independently. One participant recalled that they “*weren’t getting any work either*“, leaving them to rely on resources such as BBC Bitesize (a free online educational resource for UK students aged 3–16), (Y010), while another explained that they needed “*some kind of direction*“ because organising learning independently was itself challenging (Y015). Despite these barriers, YP consistently expressed a desire to learn. One participant explained that they “*would rather be doing something than just be sat at home*“, adding that they “*don’t mind schoolwork*“ (Y001). Another reflected:

> *“I’ve always quite loved learning… especially science… it was always once I could get into the lesson. The learning side of it… I loved it and I still really do.”* (Y019)

Parents strongly reinforced this distinction between attendance and educational engagement. Many described children who remained academically ambitious, completed schoolwork at home whenever possible and continued to value education despite being unable to attend school. One parent explained that their child *“…wants to go [to school], he’s always loved learning*“ (P027), while another described her son as “*really bright and really interested in the world*“ rather than a child who lacked motivation (P012). Some parents also recalled YP completing substantial amounts of work independently, including “*five lessons a day, five subjects a day*“ simply to avoid falling behind their peers (P018). Alongside disrupted access to education, parents described feeling sad and worried that prolonged attendance difficulties had prevented children from fulfilling their academic potential. One parent reflected that “*her learning potential is so much higher than her learning has reached… we have failed her*“ (P023), while others worried about the implications of reduced qualifications for future education, employment and life opportunities.

Together, these accounts challenge assumptions that EBSNA reflects a lack of motivation to learn. Instead, participants consistently described YP who remained eager to learn and achieve but were unable to access education because of the distress associated with attending school. The most significant educational consequence of EBSNA was therefore not diminished ambition, but the inability to realise their educational potential.

#### Social isolation and missed adolescence

Both YP and parents described EBSNA as having profound social consequences, with many YP losing friendships, becoming increasingly isolated from peers and missing important social experiences associated with adolescence.

YP described missing the everyday routines and social experiences that school had previously provided. One participant reflected that although they did not always feel they needed a large friendship group, *“not having any friends”* was *“quite difficult at times”* and left them feeling *“quite low and very bored.”* They added:

> *“I think it does leave me without some things, someone my age could do with, you know. Routine, friends, just contact with the outside world more often than going to the Co-op [supermarket] occasionally.”* (Y001)

Although home education and alternative provision were often experienced as safer and considerably less anxiety-provoking than mainstream school, participants recognised that these settings offered fewer opportunities to develop close peer relationships. One YP explained that although they met other home-educated YP, *“you don’t really have that class environment”* where *“you make good relationships”*, adding that they sometimes wished they *“could go back”* simply to have friends again (Y010).

Parents’ accounts corroborated these experiences, describing increasing social withdrawal as attendance difficulties became established. They often viewed isolation as a consequence of overwhelming anxiety, emotional exhaustion and autistic burnout rather than a preference to be alone. One parent explained that over the summer her son *“pretty much didn’t want to see anyone. He didn’t want to leave his room”* (P034), while another reflected simply that *“she wasn’t leaving the house”* (P026). Parents also worried that prolonged absence deprived their children of important developmental opportunities to build independence, friendships and social confidence.

For some YP, these social consequences persisted well beyond mainstream education. Experiences of bullying, rejection and prolonged isolation undermined confidence in forming relationships and continued to shape how they engaged with peers. One participant explained:

> *“I still struggle just to go into my own town… it’s just the fear almost kind of paralyses you… it’s harder for me to build relationships with kids my age… and it just further isolates me.”* (Y019)

Together, these accounts suggest that the consequences of EBSNA extended far beyond school attendance, disrupting friendships, limiting opportunities for social participation and restricting key developmental experiences of adolescence. For many YP, the effects of prolonged isolation persisted long after they had left mainstream education, continuing to shape their confidence, relationships and sense of belonging.

### EBSNA as a whole-family experience

Across both YP’s and parents’ accounts, EBSNA was experienced not only by the young person but by the whole family. Parents described how supporting a child experiencing EBSNA reshaped family relationships, daily routines, parental wellbeing, employment and finances, while YP recognised the emotional impact their difficulties had on those around them.

YP described the emotional demands of coping with school continuing long after they returned home. Several explained that they masked their distress while at school, only feeling safe enough to release it once they returned home, where they often became overwhelmed. One young person reflected:

> *“I’d come home and I’d just be kind of like not wanting to speak to anyone. And I’d take out my anger on my family (…) And a lot of it would pent up and then just explode and then I wouldn’t want to go the next day, or I’d go to bed late.”* (Y010)

Parents’ accounts complemented these experiences by describing the intensive emotional support that followed. Many explained that helping their child regulate emotions, recover after school and manage periods of acute distress became an everyday responsibility that dominated family life. As one parent explained:

> *“When she comes home, she is literally off the kilter and it’s quite hard to manage her. So it’s about calming her down and just trying to… there’s so much effort from our side. (…) It’s literally like a full-time job just dealing with her and trying to keep her going.”* (P019)

Parents also described how family life increasingly revolved around their child’s emotional wellbeing, often creating tension between partners and affecting siblings, whose own needs were frequently overshadowed. One parent reflected that her daughter’s distress frequently dominated family life, leaving little space for her sibling’s needs:

> *“I don’t even know how my other daughter copes with it all to be honest. [Child] was screaming down the house till 4:00 in the morning and her sister had to wake up like in two hours, and then she went and did her English final exams.”* (P019)

Others described increasing strain within family relationships as they struggled to balance supporting their child with the competing needs of other family members. One parent reflected:

> *“…there’s a lot of tension between [child] and his father. Consequently, there’s a lot of tension between me and [child]’s father, because I’m in the background going ‘you can’t do that. He’s autistic. He can’t deal with that stress.’ Our family has become essentially quite dysfunctional as a result of this experience.”* (P012)

The cumulative demands of caregiving had profound consequences for parents’ own wellbeing. Many described chronic stress, disrupted sleep, anxiety and emotional exhaustion, with several reflecting that they experienced depression for the first time while supporting their child. Others described requiring antidepressants, using alcohol to cope and feeling increasingly isolated as the demands of caregiving intensified (P020). As one parent explained:

> *“And for the first time in my life, I had a depressive episode where I was like, maybe if I just kind of fell down the stairs accidentally on purpose and had some time off in hospital, you know?”* (P034)

Alongside the emotional burden, parents described substantial practical and financial consequences. Many reduced their working hours or left employment because their child required continuous care and support, while the costs of tribunals, educational provision and reduced income created considerable financial strain. One parent summarised the impact:

> *“(…) the tribunal costs so much money to do. It also means I can’t work. Therefore, we’re forced to be on benefits to survive. We have no money whatsoever. It pushes you into poverty.” (P023)*

Many parents also described becoming increasingly socially isolated. Caring responsibilities left little opportunity to maintain employment, friendships or leisure activities, while guilt, shame and fear of judgement discouraged many from seeking support. As attendance difficulties persisted, families described their world becoming progressively smaller. One parent summarised this experience:

> *“…suddenly our world became very small because we couldn’t do the things that we were doing very sort of automatic.”* (P031)

Together, these accounts position EBSNA as a whole-family experience. Beyond its impact on school attendance, it reshaped family relationships, parental wellbeing, daily routines and financial security, demonstrating that its consequences extend well beyond the individual YP.

## Discussion

This study explored YP’s and parents’ experiences of EBSNA, conceptualising school attendance difficulties as a dynamic socioecological process that unfolded through interactions between YP’s needs and the school environment. Across both perspectives, participants described how an increasing misalignment between individual needs and school demands contributed to escalating distress, deteriorating mental health and prolonged school absence, with absence itself becoming an additional barrier to reengagement. Together, these accounts suggest that EBSNA developed gradually as psychological safety, belonging and the capacity to cope were progressively eroded, ultimately making school attendance psychologically unsustainable rather than a matter of choice.

These findings are consistent with Bronfenbrenner’s bioecological model (52) the Kids and Teens at School (KiTeS) Framework (53), and Leduc et al.’s (2024) ecological model of school refusal (11), all of which conceptualise attendance difficulties as arising through interactions between individual, family and school factors. Qualitative accounts from YP and parents in this and other studies (?) highlight EBSNA as emerging through interactions between emotional wellbeing and school environments, with relationships with teachers, school systems and educational demands shaping the development and maintenance of attendance difficulties (54, 55). Extending these findings, the present study conceptualises EBSNA as a gradual process in which an increasing misalignment between YP’s needs and school demands progressively eroded psychological safety, belonging and the capacity to cope, ultimately making school attendance psychologically impossible.

A prominent finding across both YP’s and parents’ accounts was the central role of neurodevelopmental needs interacting with school environments. Many participants described attendance difficulties arising not from neurodivergence itself, but from a mismatch between YP’s needs and school environments that failed to accommodate them, particularly in relation to sensory sensitivities, transitions and appropriate adjustments. This is consistent with recent UK evidence indicating that one in six autistic students did not attend school at all between September 2025 and early 2026 because schools were unable to meet their needs (56). Sensory overwhelm emerged as a particularly important contributor to EBSNA, adding to growing evidence that the sensory characteristics of school environments such as excessive noise, crowding, visual complexity and unpredictability create significant barriers to participation and learning for many YP with neurodevelopmental conditions (5, 32, 41, 42, 45, 57–60). These environmental demands have consistently been associated with poorer mental health, reduced participation and increased school attendance difficulties (5, 41, 61, 62). Together, these findings suggest that, for many neurodivergent YP, persistent sensory demands played a central role in escalating overwhelm, contributing to deteriorating mental health and school increasingly being experienced as an unsafe environment.

The transition to secondary school emerged as a critical turning point, when simultaneous increases in academic, organisational, sensory and social demands often transformed previously manageable challenges into sustained school attendance problems. This is consistent with qualitative and longitudinal research identifying the primary-to-secondary transition as a key period of vulnerability for EBSNA, characterised by escalating anxiety and attendance difficulties as YP negotiate unfamiliar environments, relationships and expectations (34, 54, 55). Saxton et al. (2025) further demonstrated that these challenges were amplified when schools failed to understand neurodiversity, implement appropriate adjustments or foster trusting relationships with staff (63), closely reflecting participants’ experiences in the present study.

In the current study participants across both groups consistently described psychological safety and belonging as central to school attendance, with bullying, unmet needs, punitive school practices and disrupted relationships with trusted adults gradually undermining YP’s connection to school. This aligns with qualitative evidence showing that school non-attendance develops through the cumulative impact of school-related stressors rather than a single precipitating event. Feeling misunderstood by teachers, difficult peer relationships, unmet needs and diminished school belonging are consistently identified as central to YP’s experiences of EBSNA (35, 38, 47). Teacher–pupil relationships may be particularly important, with supportive relationships associated with better mental health and problematic relationships predicting later psychiatric difficulties (64, 65).

Participants in the present study further described how anxiety, sensory overload and other expressions of distress were frequently misinterpreted as behavioural problems, resulting in punitive disciplinary responses that intensified distress and undermined trust in school. This is consistent with evidence that exclusionary and punitive behaviour management adversely affects children’s emotional wellbeing (66–69), damages relationships between schools and families (70), and disproportionately affects YP with SEND and mental health difficulties who also experience higher rates of school absence (71, 72).

The current study also highlighted the self-perpetuating nature of EBSNA. Once absence became prolonged, regardless of whether it was initially triggered by emotional distress, physical illness or other circumstances, declining confidence, loss of social connections and increasing uncertainty about returning, became additional barriers to re-engagement. This cyclical process is reflected in recent research showing that prolonged absence made returning increasingly difficult as missed learning accumulated, friendships changed and pupils became accustomed to being at home (73). Anxiety about returning further reinforced this cycle, while similar patterns have been observed among YP with chronic health conditions, where unavoidable absence increased social isolation and the demands associated with returning to school (74).

In the present study, both YP and parents consistently described a strong desire to learn, with educational disadvantage arising from an inability to access education rather than diminished academic aspirations. This challenges interpretations of non-attendance as educational disengagement and aligns with evidence that YP often remain motivated to learn and attend school despite emotional barriers that make attendance difficult (34, 54). Indeed, high academic motivation may itself create pressure, with perfectionism and fear of failure associated with greater school refusal and somatic symptoms (75, 76).

Beyond its educational consequences, EBSNA disrupted friendships and opportunities for social development, contributing to persistent isolation, while also affecting the wider family through deteriorating parental wellbeing, strained family relationships, impacts on siblings, and substantial employment and financial pressures. These findings align with previous research highlighting the social consequences of prolonged absence (77) and positioning EBSNA as a whole-family experience characterised by disruption to family functioning, parental distress and considerable practical and financial sacrifice (37, 55, 78).

### Strengths and limitations

A key strength of this study is its rigorous qualitative methodology and integration of both YP’s and parents’ perspectives, providing a richer understanding of the development and consequences of EBSNA than either perspective alone. In-depth semi-structured interviews enabled detailed exploration of participants’ experiences, while offering YP the choice of verbal or written interviews promoted inclusive participation by enabling those who did not feel able to speak with a researcher. The use of the Framework Method, independent coding by multiple researchers, regular reflexive discussions and collaborative interpretation enhanced the credibility and transparency of the findings. Development of the interview guides with the Attend Well Parent Advisory Group and YP further strengthened the relevance and acceptability of the study. Importantly, the findings contribute a novel conceptualisation of EBSNA as a dynamic socioecological process in which interactions between YP’s needs and school environments progressively erode psychological safety and belonging, ultimately making school attendance psychologically unsustainable.

Several limitations should also be acknowledged. The parent sample consisted exclusively of mothers and therefore does not capture the perspectives of fathers or other caregivers. Both samples were predominantly White British, and parents were generally highly educated, which may limit the transferability of the findings to more ethnically and socioeconomically diverse populations. Although the YP sample included participants across a range of educational settings, including mainstream, specialist and alternative provision, most participants were neurodivergent or had identified mental health difficulties, reflecting the characteristics of many YP experiencing EBSNA but potentially limiting transferability to those without SEND or mental health needs. In addition, although offering written interviews enabled participation from YP who may otherwise have been excluded, these interviews generally yielded less detailed accounts than verbal interviews. Finally, as participants were recruited through convenience sampling and were interviewed at a single time point, the findings may not reflect the experiences of all families affected by EBSNA or capture how experiences change over time.

### Implications for future research and educational practice

Our findings suggest that schools should move beyond attendance-focused interventions towards responses that prioritise psychological safety, connectedness to school, trusting relationships, and the early recognition of unmet needs. Greater understanding of neurodivergence, mental health and EBSNA may help school staff recognise distress-driven behaviours, implement appropriate adjustments and provide flexible, individualised support before attendance difficulties become entrenched. These findings are particularly relevant in the context of the recent SEND reforms, which place greater emphasis on meeting children’s needs within mainstream schools through inclusive practice, rather than relying on specialist provision, alongside effective universal and targeted support (79). Achieving this is likely to require sustained investment in staff training, resources and whole-school inclusive practices that enable schools to identify and respond to unmet needs before attendance difficulties develop.

Future research should integrate the perspectives of YP, parents, school staff and other professionals to further understand the relational and systemic processes underpinning EBSNA. Longitudinal studies following YP across the transition to secondary school would help identify the mechanisms through which attendance difficulties emerge and become entrenched, while robust evaluations of relational, trauma-informed and neuro-affirmative approaches within mainstream schools are needed to strengthen the evidence base for prevention and early intervention. Given the proposed SEND reforms and the absence of a consistent national approach to identifying and recording EBSNA, future research should also examine how changes to SEND provision influence the identification, prevalence and educational outcomes of YP experiencing attendance difficulties.

## Supporting information

Themes and subthemes

COREQ checklist

Topc guides

## Data Availability

All data produced in the present study are available upon reasonable request to the authors

## Acknowledgements

We would like to thank all the young people and parents who participated in the interviews and generously shared their experiences with us. We are also grateful to Nessie for supporting the study and participant recruitment. Finally, we would like to thank the members of our Parent and Professional Advisory Panel and Young People’s Advisory Group for their valuable guidance throughout the study and for reviewing and providing feedback on the study materials and manuscript.

## References

1. House of Commons Education Committee. Persistent absence and support for disadvantaged pupils. House of Commons.; 2023.

2. Dee TS. Higher chronic absenteeism threatens academic recovery from the COVID-19 pandemic. Proceedings of the National Academy of Sciences. 2024;121(3):e2312249121.

3. Department for Education. Pupil absence in schools in England. Academic year 2024/25. . In: Education Df, editor. May 2026.

4. Kearney CA. Framing chronic absenteeism and emotionally-based school absenteeism as public health problems. Frontiers in Child and Adolescent Psychiatry. 2025;4:1662093.

5. Gök MF, Bilgiç B, Yeşilkaya C, Turan S, Bond C. Developmental Differences in the Emotional and Behavioural Profiles of Emotionally Based School Avoidance (EBSA) in Children and Adolescents. Psychiatric Quarterly. 2026:1–13.

6. Leslie R, Oberg G, Townley C, Westphal T, Rogers L, Brömdal A. School can’t: A conceptual framework for reframing school refusal and recognising school related stress/distress. British Journal of Sociology of Education. 2025;46(7-8):1052–70.

7. Hammond-Price C, Bond C, Hatton-Corcoran S, Lyons R. An analysis of UK local authority (LA) emotionally based school avoidance (EBSA) guidance. Educational Psychology in Practice. 2025;41(3):332–52.

8. Heyne D. Developments in classification, identification, and intervention for school refusal and other attendance problems: Introduction to the special series. Cognitive and behavioral Practice. 2019;26(1):1–7.

9. Havik T, Ingul JM. How to Understand School Refusal. Frontiers in Education. 2021;6.

10. Gonzálvez C, Sanmartín R, Vicent M, García-Fernández JM. Exploring different types of school refusers through latent profile analysis and school-related stress associations. Science Progress. 2021;104(3):00368504211029464.

11. Leduc K, Tougas A-M, Robert V, Boulanger C. School refusal in youth: A systematic review of ecological factors. Child Psychiatry & Human Development. 2024;55(4):1044–62.

12. Secer I, Cimen F, Ulaş S, Ay E, Tatlı E, Garcia D, Sunay C. School refusal and its underlying mechanisms: a network perspective on individual and environmental determinants. School Mental Health. 2025;17(4):1283–99.

13. Klan A, Whitley J, Krause A, McBrearty N, Rogers MA, Smith JD. An exploration of school attendance problems experienced by children receiving mental health services. Educational & Child Psychology. 2024;41(1).

14. Devine E. An exploration of teachers’ attributions for the causes of emotionally based school avoidance. University of Nottingham; 2021.

15. Pervez A, Tattum J, McDonald C. Staff perspectives on Emotionally Based School Avoidance: a case study in a Welsh primary school. Emotional and Behavioural Difficulties. 2026:1–17.

16. Department for Education. Summary of responsibilities where a mental health issue is affecting attendance. 2023.

17. Allison MA, Attisha E, Lerner M, De Pinto CD, Beers NS, Gibson EJ, et al. The link between school attendance and good health. Pediatrics. 2019;143(2).

18. Hamilton LG. Emotionally based school avoidance in the aftermath of the COVID-19 pandemic: Neurodiversity, agency and belonging in school. Education sciences. 2024;14(2):156.

19. Lester KJ, Michelson D. Perfect storm: emotionally based school avoidance in the post-COVID-19 pandemic context. BMJ Mental Health. 2024;27(1).

20. McDonald B, Lester KJ, Michelson D. ‘She didn’t know how to go back’: School attendance problems in the context of the COVID- 19 pandemic—A multiple stakeholder qualitative study with parents and professionals. British Journal of Educational Psychology. 2023;93(1):386–401.

21. Children and young people more worried than ever about going to school. [press release]. February 2024 2024.

22. Klein M, Sosu E. School absences, academic achievement, and adolescents’ post-school destinations. Oxford Review of Education. 2025;51(3):339–56.

23. Klein M, Sosu EM, Dare S. School absenteeism and academic achievement: does the reason for absence matter? AERA open. 2022;8:23328584211071115.

24. Smerillo NE, Reynolds AJ, Temple JA, Ou S-R. Chronic absence, eighth-grade achievement, and high school attainment in the Chicago Longitudinal Study. Journal of school psychology. 2018;67:163–78.

25. Epstein S, Roberts E, Sedgwick R, Polling C, Finning K, Ford T, et al. School absenteeism as a risk factor for self-harm and suicidal ideation in children and adolescents: a systematic review and meta-analysis. European child & adolescent psychiatry. 2020;29:1175–94.

26. Epstein S, Ching BC, Wickersham A, Dutta R, Morgan C, Ford T, et al. Longitudinal associations between school absence and mental health problems in adolescents in England: a retrospective cohort study using linked administrative data. Child and Adolescent Mental Health. 2026.

27. Gonzálvez C, Inglés CJ, Kearney CA, Sanmartín R, Vicent M, García-Fernández JM. Relationship between school refusal behavior and social functioning: a cluster analysis approach. European Journal of Education and Psychology. 2019;12(1):17–29.

28. Ansari A, Hofkens TL, Pianta RC. Absenteeism in the first decade of education forecasts civic engagement and educational and socioeconomic prospects in young adulthood. Journal of Youth and Adolescence. 2020;49:1835–48.

29. Gakh M, Coughenour C, Assoumou BO, Vanderstelt M. The relationship between school absenteeism and substance use: An integrative literature review. Substance Use & Misuse. 2020;55(3):491–502.

30. Ward S, Williams J, van Ours JC. Delinquency, arrest and early school leaving. Oxford Bulletin of Economics and Statistics. 2021;83(2):411–36.

31. Finning K, Ukoumunne OC, Ford T, Danielson-Waters E, Shaw L, Romero De Jager I, et al. The association between anxiety and poor attendance at school–a systematic review. Child and adolescent mental health. 2019;24(3):205–16.

32. Fielding C, Streeter A, Riby DM, Hanley M. Neurodivergent pupils’ experiences of school distress and attendance difficulties. Neurodiversity. 2025;3:27546330251327056.

33. Halligan C, Cryer S. Emotionally based school avoidance (EBSA): Students’ views of what works in a specialist setting. Continuity in Education. 2022;3(1):13–24.

34. Slifi J. Exploring Experiences of Emotionally Based School Avoidance Across the Primary to Secondary School Transition: the Perspectives of Young People, Parents/Carers and School Staff: UCL (University College London); 2024.

35. Corcoran S, Kelly C. A meta-ethnographic understanding of children and young people’s experiences of extended school non-attendance. Journal of Research in Special Educational Needs. 2023;23(1):24–37.

36. Lissack K, Boyle C. Parent/carer views on support for children’s school non-attendance:‘How can they support you when they are the ones who report you?’. Review of Education. 2022;10(3):e3372.

37. Onslow J, Cartmell H. Understanding the parental experience of emotional barriers to school attendance: a systematic literature review. Educational Psychology in Practice. 2025;41(2):248–68.

38. Hejl C, Ezzaaf Fryland N, Hansen RB, Nielsen K, Thastum M. A review and qualitative synthesis of the voices of children, parents, and school staff with regards to school attendance problems in the Nordic countries. Scandinavian Journal of Educational Research. 2025;69(7):1490–504.

39. Saxton J, Burn A-M, Zhang X, Toulmin H, Parker J, Casey H, et al. Barriers, enablers and outcomes reported by parents engaged with the special educational needs system in England: A qualitative study. PLoS One. 2025;20(11):e0335606.

40. Wang H, Qin Q. The role of anxiety and trauma in predicting school avoidance among students: A structural equation modeling analysis. Frontiers in Psychology. 2025;16:1687797.

41. Berg AD, Sørløk K, Bretfeld-Wolf A-K, Page AG, Storås MH, Chahboun S. The missing piece in inclusion: addressing school avoidance among children with autism. Frontiers in Psychology. 2026;17:1724420.

42. van der Werff S, Bullen P, Sharp R. Understanding the contextual variables that contribute to the school refusal behavior of neurodivergent adolescents: A scoping review. Review Journal of Autism and Developmental Disorders. 2026:1–14.

43. Chian J, Holliman A, Pinto C, Waldeck D. Emotional based school avoidance: Exploring school staff and pupil perspectives on provision in mainstream schools. Educational and Child Psychology. 2024;41(1):55–75.

44. Connolly SE, Constable HL, Mullally SL. School distress and the school attendance crisis: a story dominated by neurodivergence and unmet need. Frontiers in Psychiatry. 2023;14:1237052.

45. Connolly SE, Constable HL, Mullally SL. School distress and the school attendance crisis: a story dominated by neurodivergence and unmet need. Frontiers in Psychiatry. 2023;Volume 14 - 2023.

46. Gray L, Hill V, Pellicano E. “He’s shouting so loud but nobody’s hearing him”: A multi-informant study of autistic pupils’ experiences of school non-attendance and exclusion. Autism & Developmental Language Impairments. 2023;8:23969415231207816.

47. Møller G. Early identification of school refusal from parents’ perspectives. Education Sciences. 2025;15(9):1211.

48. Gale NK, Heath G, Cameron E, Rashid S, Redwood S. Using the framework method for the analysis of qualitative data in multi-disciplinary health research. BMC medical research methodology. 2013;13(1):117.

49. Ritchie J, Ormston R, McNaughton Nicholls C, Lewis J. Qualitative research practice: A guide for social science students and researchers. 2013.

50. Lumivero. NVivo (version 15). 15 ed2025.

51. Tong A, Sainsbury P, Craig J. Consolidated criteria for reporting qualitative research (COREQ): a 32-item checklist for interviews and focus groups. International journal for quality in health care. 2007;19(6):349–57.

52. Bronfenbrenner U, Ceci SJ. Nature-nurture reconceptualized in developmental perspective: a bioecological model. Psychological review. 1994;101(4):568.

53. Melvin GA, Heyne D, Gray KM, Hastings RP, Totsika V, Tonge BJ, Freeman MM, editors. The Kids and Teens at School (KiTeS) framework: An inclusive bioecological systems approach to understanding school absenteeism and school attendance problems. Frontiers in education; 2019: Frontiers Media SA.

54. Musenga-Grant L, MacFarland K, White A. Exploring the voices of children and young people in relation to emotionally based school non-attendance. Educational & Child Psychology. 2025;42(2):5.

55. Want H, Gulliford A. Barriers to school attendance as experienced by young people and their parents: A Narrative Oriented Inquiry. Educational and Child Psychology. 2024;41(1):9–30.

56. Ambitious about Autism. When will we learn: Lost Learning report.: Ambitious about Autism; 2025.

57. Al Qutub R, Luo Z, Vasilikou C, Tavassoli T, Essah E, Marcham H. Impacts of school environment quality on autistic pupil’s behaviours – A systematic review. Building and Environment. 2024;265:111981.

58. Birkett L, McGrath L, Tucker I. Muting, filtering and transforming space: Autistic children’s sensory ‘tactics’ for navigating mainstream school space following transition to secondary school. Emotion, Space and Society. 2022;42:100872.

59. Havik T, Bru E, Ertesvåg SK. Parental perspectives of the role of school factors in school refusal. Emotional and behavioural difficulties. 2014;19(2):131–53.

60. Price J, Romualdez AM. ‘It just feels unnatural being here’: Autistic secondary school students’ experiences of sensory sensitivities in the school environment. Autism. 2025;29(9):2228–38.

61. Block SM. Classroom design and its influence on students’ performance within the autism spectrum diagnosis. 2018.

62. Martin R, Wilkins J. Creating visually appropriate classroom environments for students with autism spectrum disorder. Intervention in School and Clinic. 2022;57(3):176–81.

63. Saxton J, Matthews J, Winterburn I, Casey H, Zylbersztejn A, Barnes S, et al., editors. Exploring the experiences and outcomes of children and young people receiving support for special educational needs over time in England: a qualitative study. Frontiers in Education; 2025: Frontiers Media SA.

64. Marlow R, Goodman R, Meltzer H, Ford T. Influence of problematic child-teacher relationships on futurepsychiatric disorder: population survey with 3-yearfollow-up. The British Journal of Psychiatry. 2013;202(5):336–41.

65. Severyn M, Ford TJ. 6.38 A Cross-Sectional Analysis Assessing the Association Between Problematic Teacher-Pupil Relationships and Psychiatric Disorder, As Reported By Parents and Children. Journal of the American Academy of Child & Adolescent Psychiatry. 2024;63(10):S293.

66. Barker J, Alldred P, Watts M, Dodman H. Pupils or prisoners? Institutional geographies and internal exclusion in UK secondary schools. Area. 2010;42(3):378–86.

67. Christie C, Harding E. Restrictive practices in English schools: an interpretative phenomenological analysis of parental lived experience. Frontiers in Education. 2025;Volume 10 - 2025.

68. Condliffe E. ‘Out of sight, out of mind’: an interpretative phenomenological analysis of young people’s experience of isolation rooms/booths in UK mainstream secondary schools. Emotional and Behavioural Difficulties. 2023;28(2-3):129–44.

69. Sealy J, Abrams EJ, Cockburn T. Students’ experience of isolation room punishment in UK mainstream education.‘I can’t put into words what you felt like, almost a dog in a cage’. International Journal of Inclusive Education. 2023;27(12):1336–50.

70. Trippler K, Anderson J, Burn A-M. Internal exclusion as a “dumping ground”: analysis of the perspectives of parents whose children have experienced internal exclusion in UK schools. Frontiers in Education. 2025;Volume 10 - 2025.

71. Parker C, Paget A, Ford T, Gwernan-Jones R. ‘. he was excluded for the kind of behaviour that we thought he needed support with…’A qualitative analysis of the experiences and perspectives of parents whose children have been excluded from school. Emotional and Behavioural Difficulties. 2016;21(1):133–51.

72. Tejerina-Arreal M, Parker C, Paget A, Henley W, Logan S, Emond A, Ford T. Child and adolescent mental health trajectories in relation to exclusion from school from the Avon Longitudinal Study of Parents and Children. Child and Adolescent Mental Health. 2020;25(4):217–23.

73. Moore E, Walker M. Voices from the Classroom: Understanding How Secondary Schools Support Pupils Returning from Absence. National Foundation for Educational Research. 2025.

74. Herlitz L, Jay MA, Powell C, Deighton J, Gilbert R, Blackburn R, editors. Challenging school experiences of unavoidable absence and return for young people with chronic health conditions in English secondary schools: a qualitative, multi-informant study. Frontiers in Education; 2025: Frontiers Media SA.

75. Fuster A, Vicent M, Pérez-Marco M, Gonzálvez C. Perfectionism and School Non-Attendance Reasons in Adolescents: A Latent Profile Analysis: Perfectionism and School Non-Attendance. Journal of Adolescence. 2026;98(5):1681–91.

76. Pérez-Marco M, Gonzálvez C, Fuster A, Vicent M. Identifying Functional Profiles of School Refusal and Perfectionism in Adolescents. European Journal of Education. 2025;60(3):e70201.

77. Tamlyn G. An exploration of the narratives of young people who have experienced Emotional Based School Avoidance: University of East London; 2022.

78. Shah ZT. Exploring parental experiences of emotionally based school avoidance: using interpretative phenomenological analysis: University of Birmingham; 2024.

79. Department for Education. Every child achieving and thriving. 2026.

