## Supplementary material for "“It was like a weird, paralysing thing”: Understanding the development and consequences of emotionally based school non-attendance (EBSNA) through young people’s and parents’ experiences": Themes and subthemes

### Appendix 3: Final thematic framework: themes, subthemes and descriptions

| **Theme** | **Subtheme** | **Description** |
| --- | --- | --- |
| **1. Misalignment between needs and the school environment** | **Individual needs interacting with school demands** | EBSNA developed through an increasing mismatch between YP’s individual needs and the demands of the school environment. Emotional, physical and neurodevelopmental differences increased vulnerability when environments did not accommodate needs, including sensory sensitivities, need for predictability and difficulties managing organisational and social demands. |
|  | **Transition to secondary school as a turning point** | The move to secondary school often intensified previously manageable difficulties through simultaneous increases in academic, organisational, sensory and social demands, alongside loss of familiar routines, relationships and environments. Inadequate transition planning and poor transfer of information about needs and adjustments could further compound these difficulties. |
|  | **Losing a sense of safety and belonging** | Repeated experiences of bullying and social exclusion, feeling misunderstood by staff, unmet needs, inconsistent relationships with trusted adults and punitive or inflexible school practices progressively undermined YP’s psychological safety, trust and sense of belonging at school. |
| **2. Becoming unable to attend school** | **When coping became unsustainable** | Escalating anxiety and the sustained effort required to cope with school progressively exceeded YP’s psychological resources. Distress increasingly extended beyond specific school situations to anticipation of school itself and was accompanied by deterioration in mental health and functioning, sometimes culminating in withdrawal, burnout, self-harm or acute mental health crisis. |
|  | **Absence becomes a barrier to returning** | Once absence became prolonged, it created additional barriers to re-engagement. Loss of routine, confidence and friendships, falling behind academically, fear of judgement and increasing unfamiliarity with school made returning progressively more difficult, sometimes overshadowing the factors that had initially contributed to absence. |
| **3. The wider impact of EBSNA** | **Disrupted education despite desire to learn** | EBSNA disrupted access to learning and limited opportunities to fulfil academic potential, often with little structured educational provision during absence. YP nevertheless commonly remained motivated to learn and achieve, indicating that disrupted education reflected difficulty accessing school rather than educational disengagement. |
|  | **Social isolation and missed adolescence** | Prolonged attendance difficulties disrupted friendships, everyday contact with peers and opportunities to participate in social and developmental experiences associated with adolescence. Isolation could persist beyond mainstream schooling and affect YP’s confidence, relationships and sense of belonging. |
|  | **EBSNA as a whole-family experience** | EBSNA affected the wider family as parents took on intensive emotional and practical caregiving responsibilities. Consequences included poorer parental wellbeing, disrupted family relationships and routines, impacts on siblings, reduced employment and income, financial pressures and increasing social isolation. |
