## Supplementary material for "“It was like a weird, paralysing thing”: Understanding the development and consequences of emotionally based school non-attendance (EBSNA) through young people’s and parents’ experiences": Topc guides

### Appendix 1: Parent and Young Person Interview Topic Guides

##### Attend Well: Parent/carer interview topic guide

Structure: Semi-structured interview lasting up to 60 minutes, either in person or online (depending on preference).

Objective: To understand the reasons behind persistent absence and what needs to be changed and by whom to help bring absent students back to classroom.

#### Pre-Interview checks: Participants have read and understood the Participant Information Sheet and signed a consent form

#### Introduction:

- Researcher will begin by introducing themselves thanking participant for agreeing to take part in the study.
- Explain how the interview will run and how long it will take.
- Remind participant that the interview will be recorded and remind them that the answers and information they share will be held confidentially and will not be passed on to family members or professionals (except if safeguarding issues are raised, explain clearly what this may involve, what would happen and that the participant will be involved in discussions or decisions).

#### Consent:

- Ask the participants individually if they have any questions and answer any that they might have.
- Ensure a consent form has been signed by both researcher and participant, and that participant received a copy.
- Remind participant that they can chose not to answer some questions or stop the interview at any time without giving a reason.
- Ask participant’s permission to audio record the interview. If the interview is online, ask whether they prefer their camera on or off, and if it’s on, ask permission to record both audio and video, and delete video recording immediately after the interview is completed.

##### Interview Topic Guide

**NB:** This topic guide reflects the nature of the questions that are expected to be asked during the interview. It should be acknowledged that this is not a script, instead these questions are expected to prompt a natural discussion between researcher and young participant. Each question will be asked/framed in a way which is appropriately tailored to participant’s age. At all times, questions and discussion points will be phrased in a sensitive manner and accordingly to the context.

#### Background questions:

- How many children do you have? How old are they?
- Do they all go to the same school?

#### School attendance:

- Does your child go to school full time? *(Example follow-up questions: On average, how many hours are they in school every day? Are they in any other alternative type of education?)*
- When going to school became difficult for them?
- Do you think young person’s mental health has impact on their school attendance?

#### Your child’s school:

- Is/was there anything that you really liked about your child’s school? *(Example prompts: a teacher, best friend, especially interesting subject)*
- Is/was there anything that you really disliked about your child’s school?
- Do you remember why going to school became difficult for them? *(Example follow-up questions: Was it a specific event or a number of small things that added up?)*

#### Support:

- Has your child received any support to help them stay at school/go more often? *(Example follow-up questions: When did they get the support (when the problem started or later on)? Who provided the support?)*
- How easy/difficult was finding support? *(Example follow-up questions: Has anyone signposted you?*
- If yes, was it helpful? *(Example follow-up questions: What was especially helpful/unhelpful?)*

#### Going back to school:

- What do you think would have to change for your child to feel happy and safe in school?
- What support do you think they need to go back to school? *(Example follow-up questions: Who should provide it?)*

#### Closing:

- Thank participant for their time and contributions.
- Ask if there’s anything they would like to add that hasn’t been discussed.
- Inform participant that they will receive a thank you voucher via email or post (as per their preferred option)
- Ask if they would like to receive a summary report of the key findings.
- Ask if they have any questions

### Attend Well: Young Person interview topic guide

Structure: Semi-structured interview lasting up to 60 minutes, either in person or online (depending on preference).

Objective: To understand the reasons behind persistent absence and what needs to be changed and by whom to help bring absent students back to classroom.

#### Pre-Interview checks: (1) Participants have read and understood the Participant Information Sheet. (2) Researcher has obtained Informed Consent from young people aged 16+. For those aged under the age of 16 years, assent has been obtained, as well as parental consent.

#### Introduction:

- Researcher will begin by introducing themselves thanking participant for agreeing to take part in the study.
- Explain how the interview will run and how long it will take.
- Remind participant that the interview will be recorded and remind them that the answers and information they share will be held confidentially and will not be passed on to family members or professionals (except if safeguarding issues are raised, explain clearly what this may involve, what would happen and that the participant will be involved in discussions or decisions).

#### Consent:

- Ask participant if they have any questions and answer any that they might have.
- Ensure a consent form has been signed by both researcher and participant, and that participant received a copy.
- Remind participant that they can chose not to answer some questions or stop the interview at any time without giving a reason.
- Ask participant’s permission to audio record the interview. If the interview is online, ask whether they prefer their camera on or off, and if it’s on, ask permission to record both audio and video, and delete video recording immediately after the interview is completed.

### Interview Topic Guide

**NB:** This topic guide reflects the nature of the questions that are expected to be asked during the interview. It should be acknowledged that this is not a script, instead these questions are expected to prompt a natural discussion between researcher and young participant. Each question will be asked/framed in a way which is appropriately tailored to participant’s age. At all times, questions and discussion points will be phrased in a sensitive manner and accordingly to the context.

#### Warm up questions:

- What is your favourite TV show and why do you like it?
- What was the funniest thing you’ve done when you were little?

#### Your school:

- Is/was there anything that you really liked about your school? *(Example prompts: a teacher, best friend, especially interesting subject)*
- Is/was there anything that you really disliked about your school?

#### School attendance:

- Do you currently go to school full time? *(Example follow-up questions: On average, how many hours you are in school every day? Are you in any other alternative type of education?)*
- When going to school became difficult for you?
- Do you remember why going to school became difficult for you? *(Example follow-up questions: Was it a specific event or a number of small things that added up?)*
- Do you think there is a link between mental health and being able to go to school?

#### Support:

- Have you received any support to help you stay at school/go more often? *(Example follow-up questions: When did you get the support (when the problem started or later on)? Who provided the support?)*
- If yes, was it helpful? *(Example follow-up questions: What was especially helpful/unhelpful?)*

#### Going back to school:

- What would have to change for you to feel happy and safe in school?
- What support would you like to get to help you go back to school? *(Example follow-up questions: Who should provide it?)*
- What an ideal school look like for you? *(Example prompts: tell me about an ideal school day, teachers, friends.)*

#### Closing:

- Thank participant for their time and contributions.
- Ask if there’s anything they would like to add that hasn’t been discussed.
- Inform participant that they will receive a thank you voucher via email or post (as per their preferred option)
- Ask if they would like to receive a summary report of the key findings and if they have any questions.

### Attend Well: Young Person online interview questions

*NB: Before taking part in a written interview, all young people received a Participant Information Sheet with detailed information about the study and procedures and were asked to provide written consent. For young people under 16, parental consent was also obtained.*

***This study is hoping to understand why some young people find it difficult to attend school and what can be done to help young people feel happier, safer and more supported in school.***

**In the space below, please tell us more about the difficulties that you have had attending school.** This might include:

- When it became difficult to go to school
- Whether you are still in school now
- How often to you go to school

**We would like to understand more about why some young people can find it difficult to attend school.** We know that for some young people, it might be difficulties with:

- friendships
- teachers
- rules
- busy corridors
- noise and lights
- travel to school
- physical health
- mental health
- caring responsibilities
- and many more...

**Do you remember why going to school became difficult for you? Can you tell us a bit about the things that were/are difficult for you?**

**Have you received any support to help you stay at school/go more often?** If yes, you might also like to tell us:

- Did you find the support helpful?
- Did you get support when you started to find school difficult, or later on?

**If you are now better able to attend school, what has helped you to be able to do this?** This might include things like:

- Moving to a new setting, in which case what is better about it?
- Changes to your school environment
- Support from particular people (what did this support look like?)

**Can you tell us about your ideal school day?** You might like to think about your journey to school, spaces to sit, subjects and friends/teachers.

**Is there anything that would have to change for you to feel happy and safe in school?**

**Is there anything else that you would like to share with us that would help schools to better understand young people and why attending school can be very difficult?**

### Appendix 2: Final thematic framework: themes, subthemes and descriptions
